# Using conditional inference to quantify interaction effects of socio-demographic covariates of US COVID-19 vaccine hesitancy

**DOI:** 10.1101/2021.10.02.21264456

**Authors:** Ke Shen, Mayank Kejriwal

## Abstract

COVID-19 vaccine hesitancy has become a major issue in the U.S. as vaccine supply has outstripped demand and vaccination rates slow down. At least one recent global survey has sought to study the covariates of vaccine acceptance, but an inferential model that makes simultaneous use of several socio-demographic variables has been lacking. In this article, we present such a model using US-based survey data collected by Gallup. Our study agrees with the global survey results in some respects, but is also found to exhibit significant differences. For example, women and people aged between 25-54 were found to be more vaccine hesitant. Our conditional inference tree model suggests that trust in government, age and ethnicity are the most important covariates for predicting vaccine hesitancy, and can interact in ways that make them useful for communication-based outreach, especially if conjoined with census data. In particular, we found that the most vaccine hesitant individuals were those who identified as Black Republicans with a high school (or lower) education and lower income levels, who were involuntarily unemployed and trusted in the Trump administration.

## Introduction

COVID-19 vaccines were approved in late 2020 and early 2021 for public use in countries across the world. In May, President Biden announced his goal as getting at least 70% of Americans partially vaccinated against COVID-19 by early July at the latest. Current statistics indicate that this goal has not been achieved, not due to supply constraints, but rather, due to vaccine hesitancy among certain segments of the population. As shown in the latest news^1, 2^, 31% Americans are still hesitant about receiving the COVID-19 vaccine. In order to speed up vaccination and reduce hesitancy, the use of material incentives and communication-based outreach are being explored. While laudable, for maximal effectiveness, we argue that such efforts need to carefully identify vaccine-hesitant subgroups in the U.S. and investigate the most important predictors of vaccine hesitancy. For such identification to be accurate and fine-grained, while still preserving privacy, it is important to understand the interaction and *conditional* effects between relevant socio-demographic correlates that are known to be important predictors^3^. An example of a conditional effect is captured in a (illustrative-only) statement such as ‘People between 18-25 tend to be vaccine hesitant, but the hesitancy disappears if the education exceeds college level.’

Several studies have highlighted key factors associated with vaccine hesitancy in different countries, and considerable heterogeneity has been observed. Lazarus et al.^4^ provided a global survey of COVID-vaccine acceptance, with the study suggesting that men were less likely to get vaccinated than women. People with higher incomes and education were also more vaccine accepting. However, research into vaccine hesitancy in the UK^5^ suggested higher vaccine hesitancy among women, younger age groups and those with lower education levels. Minority groups, including Black individuals, were also found to be more hesitant. Similar results were shown for Portugal^6^, including individuals who had lost income during the pandemic and had lower confidence in the vaccine. In Arabic countries, researchers^7^ reported that male respondents with higher educational levels, and those with histories of chronic disease, were more COVID-vaccine accepting.

Vaccine hesitancy has been documented as a challenge several times in previous research^2, 8–10^. Coustasse, Kimble and Maxik^8^ revealed that layperson skepticism may have been compounded by political influences during the Trump presidency. Callaghan et al.^2^ explained that the two most cited reasons for vaccine hesitancy are concerns about vaccine safety, as well as lack of trust in vaccine effectiveness. Some researchers have also focused on special groups of people, such as medical students^11^ and healthcare workers^12^. Differences in ethnicity have been frequently mentioned in vaccine hesitancy research^9, 13^. In work resembling this study, multiple regression analyses have been used^9^ to caution the government into paying more attention to vaccine hesitant groups, who (in the US, at least) are often minority groups who have had negative experiences with the healthcare system in the past, including African-Americans and Hispanics. Individuals with conservative political affiliations (including Republicans in the US), and those with children at home, have also been found to be vaccine hesitant.

To facilitate comparisons with the survey results by Lazarus et al.^4^, we use univariate regressions and odds ratios on Gallup data, but to derive conditional effects, we also use an established statistical model called a conditional inference tree (CIT)^14^. CITs embed tree-structured regression models into a well-defined theory of conditional inference procedures. This technique is suitable for non-parametric data (e.g., nominal and binary), including nominal variables with many levels (as are often observed with *ethnicity* and *employment status* variables in survey-based studies such as this one). The classification tree algorithm uses a unified unbiased splitting technique (called permutation-based significance tests^15^) based on p-values for variable selection and pruning, making it more statistically robust than classification and regression trees (CARTs). Compared to logistic regression models, CITs provide us more interpretability and flexibility, while still maintaining high prediction accuracy. It has been applied in a similar context in the past to study conditional effects with respect to climate change skepticism^16^.

## Methods

### Survey participants and data collection

Gallup began fielding on March 13, 2020 by launching a specific survey that collected people’s responses during the COVID-19 pandemic, polling daily random samples of the Gallup Panel (a probability-based, nationally representative panel of U.S. adults). Women comprised 45.5% of the study population. 50.7% of all participants were in a household that earned more than 90,000$ per year. About 91% had more than a high school education, and 58.4% of the respondents were in an age range of 25-64.

As in other Gallup surveys, such as the Gallup World Poll, questions measuring employment, health, and demographics are included. The vaccine acceptance question was asked starting from July 20, 2020: ‘If an FDA-approved vaccine to prevent coronavirus/COVID-19 was available right now at no cost, would you agree to be vaccinated?’ with possible response of ‘Yes’ or ‘No’ to investigate a person’s willingness to be vaccinated. The ‘vaccine question’ was added on July 20, the responses of interest in this study are from July 20, 2020 to Jan 1, 2021, and, there are 29,399 valid responses collected across this time period. Since there were questionnaire changes (i.e., some new questions were added and some old questions were removed in an update of the survey), some participants were not asked to answer some questions. For example, the vaccine acceptance question was asked from July 20, 2020 to Jan 1, 2021, and the trust-in-government question was asked from July 20, 2020 to Sep. 14, 2020; hence, responses for the trust-in-government question after Sep. 14, 2020 will be NaN. The trust-in-government question imports 11,783 NaN values in those 29,399 responses. Similarly, responses such as ‘Not Applicable/Don’t know/Does not apply’ are also treated as NaN. After filtering records that include NaN in one or more values, 16,322 instances were left. A full statistical profile of respondents’ answers per covariate level is reproduced in Supplementary Material Table 1.

### Analysis

We analyzed the distribution of the vaccination responses against demographics and other covariates of interest for all 16,322 instances in the dataset. We computed a set of univariate logistic regressions, defining the label as 1 if a respondent answered ‘Yes’ in response to the vaccine question, and 0 if the answer was ‘No’. The vaccine acceptance before removing NaN values was measured to be 70.6%, and after NaN filtering, as described above, the vaccine acceptance remained at similar levels (71.17%), consistent with what has been reported in the press and in other US-based surveys. The independent demographic variables included age, gender, ethnic, party, employment status, income and education. A summary of valid responses to the survey questions and also the characteristics of these respondents are provided in the supplementary material. Men comprised 54.48% of the study population, over 70.7% participants came from the households which earned more than $60,000 before taxes per year, and 42.47% participants were employed full-time. White individuals constitute the majority (87%) of respondents. Around 90% of respondents obtained at least a high school diploma, and 41.12% of respondents were over 65 years old. Democrats were 41.80% of participants, and Republicans were 25.15% (the remainder were independents). Responses to the trust-in-government question (‘Please think about the recent impact of the coronavirus (COVID-19) on your life when responding to the following and indicate your level of agreement or disagreement: I have confidence in the leadership of President Donald Trump to successfully manage emerging health challenges’) were recorded on a five-point scale, from strongly disagree (1) to strongly agree (5). Values on this scale greater than a threshold of 3 were used to separate individuals who trusted in the government versus those who did not. 34.94% of respondents reported high levels of trust in the Trump government.

We then explore and analyze vaccine acceptance using a *classification conditional inference tree (CIT) model* (which uses unbiased recursive partitioning) at the individual level. CITs embed tree-structured regression models into a well-defined theory of conditional inference procedures. It is unnecessary to consider data imputation techniques, since the models generate high prediction performance (namely, accuracy of 75.5%). The R package *party* is used for our recursive partitioning analysis, with the max depth set to 4, and with the p-value required to be less than 0.01, in order for a node to be split by the model.

## Results

In response to the vaccine question, 71.2% of respondents (11,616 of 16,322) would agree to be vaccinated, or have already received a COVID-19 vaccine. Of these 11,616 individuals, 5,096 women and 6,520 men were willing to be freely vaccinated. Comparing White individuals to minorities, 72.4% White individuals had positive responses, compared to 62.2% non-White respondents. Table 1 reports results for univariate regressions for positive responses to the vaccine question using eight variables, including whether or not the respondent had confidence in the leadership of the (then Trump) administration to successfully manage emerging health challenges.

**Table 1.** Univariate regression outputs for vaccine acceptance against demographic variables and other covariates of interest. For each covariate, we show its odds ratio and 95% confidence interval.

|  | **Beta-coefficients of vaccine acceptance (95% CIs)** |
| --- | --- |
| Annual house-hold income | [36,000, 60,000) v.s. [0, 36,000) 1.09 (0.96, 1.23) |
|  | [60,000, 90,000) v.s. [0, 36,000) 1.16 (1.03, 1.31) |
|  | [90,000, 120,000) v.s. [0, 36,000) 1.20 (1.06, 1.35) |
|  | [120,000, 180,000) v.s. [0, 36,000) 1.38 (1.22, 1.56) |
|  | [180,000, 240,000) v.s. [0, 36,000) 1.61 (1.37, 1.90) |
|  | [240,000, ∞) v.s. [0, 36,000) 1.96 (1.66, 2.31) |
| Age | 25–54 v.s. 18–24, 0.55 (0.32, 0.93) |
|  | 55–64 v.s. 18–24, 0.57 (0.33, 0.96) |
|  | 65+ v.s. 18–24, 1.00 (0.58, 1.67) |
| Race/Ethnicity | Hispanic v.s. White, 0.74 (0.64, 0.86) |
|  | Black v.s. White, 0.42 (0.36, 0.49) |
|  | Asian v.s. White, 1.20 (0.91, 1.59) |
|  | Other v.s. White, 0.63 (0.47, 0.86) |
| Political Party | Republican v.s. Democrat, 0.22 (0.20, 0.24) |
|  | Independent v.s. Democrat, 0.37 (0.33, 0.40) |
|  | Other v.s. Democrat, 0.19 (0.15, 0.23) |
| Employment Status | Part-time v.s. Full-time, 0.98 (0.87, 1.11) |
|  | Involuntary unemp. v.s. Full-time, 0.88 (0.74, 1.04) |
|  | Not in labor force v.s. Full-time, 1.41 (1.31, 1.52) |
| Gender | Male v.s. Female 1.26 (1.18, 1.35)) |
| Education | High school graduate v.s. Less than a high school diploma, 2.15 (1.27, 3.65) |
|  | Technical, trade, vocational or business school or program after high school v.s. Less than a high school diploma, 1.82 (1.06, 3.10) |
|  | Some college, college, university, or community college – but no degree v.s. Less than a high school diploma, 2.43 (1.44, 4.09) |
|  | Four year bachelor’s degree from a college or university v.s. Less than a high school diploma, 3.77 (2.24, 6.35) |
|  | Some postgraduate or professional schooling after graduating college, but no post v.s. Less than a high school diploma, 4.26 (2.50, 7.25) |
|  | Two year associate degree from a college, university, or community college v.s. Less than a high school diploma, 2.60 (1.54, 4.40) |
|  | Postgraduate or professional degree, including master’s, doctorate, medical, or law v.s. Less than a high school diploma, 5.99 (3.55, 10.10) |
| Trust in government | yes v.s. no 0.23 (0.22, 0.25) |

We find that people aged 25-65 are far less likely to accept the vaccine than those aged 18-24. People aged over 65 are as willing to be vaccinated as youth (18-24), with odds ratio (OR) of 1.0 and 95% confidence interval (CI) (0.58, 1.67). Gender differences, however, were much more significant. This is different from data was reported in the worldwide survey by Lazarus et al., where people aged 25-65 were found to be more vaccine accepting than youth, and people aged 65+ were more vaccine accepting than youth as well. In other words, within the United States, older people are more vaccine hesitant relative to youth (compared to global data collected earlier last year); and worryingly, the middle spectrum of the age distribution is (or has become) much more vaccine hesitant compared to global counterparts.

The data suggests that men are more vaccine-accepting than women, with an OR of 1.26 (95% CI (1.18, 1.35)), which contrasts with the global survey findings. Concerning ethnicity, we find that, except for Asians, minorities, including Hispanic and Black individuals, are less likely to accept vaccination compared with White individuals. Black individuals, in particular, are vaccine hesitant (OR = 0.42; 95% CI (0.36, 0.49), compared to White). These findings are consistent with those reported in major press outlets in recent months.

An obvious trend can be seen with annual household incomes: compared to the lowest income group, people at higher household income levels are more likely to be vaccine accepting and the strongest difference (OR = 1.96; 95% CI (1.66, 2.31)) is observed in participants with over $240,000 household income per year, as compared to those making (at most) $36,000 annual household income. Higher levels of education were also associated positively with vaccine acceptability. An OR of 5.99 (95% CI (3.55, 10.10)) was observed for postgraduate participants relative to those with less than a high school education. People not in the labor force (full time students, retired people, and those who are unemployed, but not looking for a job) are also more likely to accept the vaccine (OR=1.41, 95% CI (1.31, 1.52)), which is consistent with the odds ratio of 65+ participants’ vaccine acceptance (compared to people in the 18-24 age range), since many individuals not in the labor force would, in all likelihood, be categorized into these two age groups. However, we also found that people with a part-time job, or with an involuntary unemployed status, are less likely to positively respond to vaccination. This is a potential cause for concern, since employer mandates or incentives would not have the impact needed to sway this group. Additionally, it is less clear if vaccine hesitancy is due to these individuals’ being too busy working or looking for a job, or due to skepticism.

Concerning political affiliation, Democrats are most inclined to vaccinate, while Republicans, Independents and also those from the third party are more likely to negatively respond to the vaccine question. Those who believed that the Trump government could successfully manage emerging health challenges were less willing to be vaccinated, compared to those who doubted the administration’s COVID-19 policies (OR=0.23, 95% CI (0.22, 0.25)). These sentiments may have shifted since the new administration was elected, which a follow-up study could aim to quantify.

Since variables like the annual household income, gender, and political party play important roles in shaping individual-level acceptability of COVID-19 vaccination, an important goal of this article is to identify the key *predictors* of vaccination willingness. The CIT, a non-parametric, unbiased recursive partitioning method, is an effective way to overcome the limitations of conventional regression methods, such as logistic regression. Our tree model achieved high classification accuracy≥(75.5%), and is parsimonious, as shown in Fig 1. As illustrated therein, the trust in the Trump administration, age, and ethnicity are the most important predictors. Only 34.9% participants had confidence that the Trump administration could successfully handle the pandemic. Again, consistent with OR shown in Table 1, those that trust in the Trump administration are less likely to be vaccinated, and the younger female Trump supporters are the least likely to be vaccinated. However, older supporters with higher education or income responded more positively to the vaccine question.

**Figure 1.**
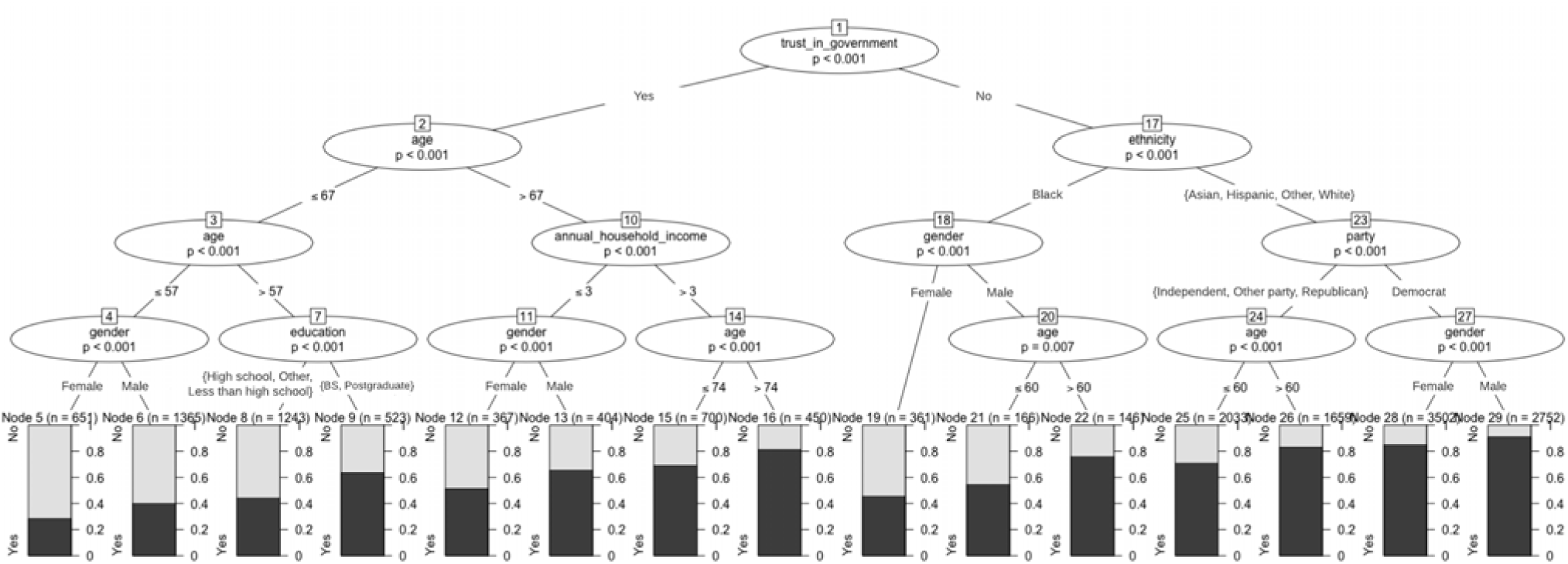
The conditional inference tree model for predicting vaccine acceptance. The education node shows the highest academic degree obtained by participants, with ‘BS’ representing a four year bachelor’s degree from a college or university. The tree is four levels deep and shows only statistically significant variables. Total sample size (N) for vaccine acceptance is 16,322.

For participants who did not trust in the Trump administration, *ethnicity* is the strongest predictor. Black individuals were less likely to respond positively compared to White, Hispanic, Asian, and other ethnic individuals. Within the Black community, there are gender-specific differences. Men are predicted to be more accepting of the vaccine than women. A similar conclusion is also obtained for other ethnicities, but party, rather than gender, is a stronger predictor. Democratic participants were significantly different from others, since more than 85% stated that they would agree to be vaccinated. Independents and Republicans were less positive, and again, compared to the older age-group, younger Independents and Republicans were less likely to say ‘yes’ to the vaccine question. We also experimented with some subjective well-being variables in the CIT model, but they were not found to play significant roles in predicting vaccine acceptability.

## Discussion

The results in the previous section illustrate considerable response diversity among different groups. Similar to Asian nations’ high vaccination acceptance shown in other work^4^, Asians in the U.S. are also the ethnic group most open to COVID-19 vaccination. There is also a relatively high tendency to accept among people with higher education and income. This is also consistent with global survey results^4^, but with one significant difference: people in the 25-64 age group are observed to be more vaccine accepting (compared to those aged 18-24), the vaccine acceptance of those aged 25-64 in our sample is smaller compared to younger people. Gender-specific differences are also observed: women tend to give positive responses to the vaccine question than men in the global study, while in our study, women responded less positively compared tomen.

The *sum of odds ratios* gives us a descriptive score for each respondent’s vaccination willingness. To be specific, the value of the *anchor choice* in each survey question is set to 1 (for example ‘White’ in Race/Ethnicity and ‘full-time’ in Employment Status); values of all other choices for a given survey question are represented by their odds ratios when regressed against the corresponding anchor choice (for example, 0.74 for ‘Hispanic’ in Race/Ethnicity; see Table 1). The sum of these odds ratios for a given individual suggests that the higher the score, the higher the odds of the individual accepting COVID-19 vaccination. Based on this methodology and our data, we can compose profiles of those most, and least, willing to be vaccinated. The former profile (i.e., who have the highest sum of odds ratios) turns out to be a male Democrat, more than 65 years old, who has a postgraduate degree, holds a full-time job or is not in the labor force (e.g., retired), has over $240,000 annual household income, and did not trust in the Trump government. The top 3 profiles correspond to such an individual who is of Asian, White and Hispanic ethnicity, respectively (in descending order).

We found 14 respondents in our survey who fit one of these top 3 profiles, and we verified from their actual responses to the vaccine question that all of them were indeed willing to be vaccinated. We also find the top 3 profiles who are the least likely to be vaccinated (i.e., those who have the lowest sum of odds ratios). The profile with the least sum-of-odds-ratios is: a Black female Republican in the 25-54 age group, who has a part-time job with gross household income less than $60,000, and with less than a high school education. The next profile is the same as above, but the individual is White instead of Black. The third profile is the same as the first profile, except that the individual has a full-time job and is engaged in a technical, trade, vocational or business school or program after high school.

Only 3 people were found to fit these (most vaccine hesitant) profiles, and none of them was willing to be vaccinated based on their actual response to the vaccine question. Additionally, in checking the top-20 profiles for the most, and the least, vaccine accepting people per the sum-of-the-odds-ratio methodology, we found that 93% (266 out of 284) high-scoring respondents responded ‘yes’ to the vaccine question, and 73% (30 out of 41) low-scoring participants responded ‘no’. Therefore, while not perfect, the odds-ratios score, in conjunction with the CIT, might be a useful tool for predicting how likely a respondent is to be vaccinated, and may help local, state and federal governments devise more effective communication and incentive strategies for meeting federal vaccination goals, which are currently falling short (at least, as of July).

## Supporting information

IRB abstract and title

IRB submitted protocol

IRB exempt approval notice

## Data Availability

All primary data used in this paper are licensed by Gallup as part of their Franklin Templeton COVID-19 survey. A statistical description of participants and breakdown of the COVID-19 vaccine acceptance questions is tabulated in the supplementary material.

https://news.gallup.com/opinion/gallup/308126/roundup-gallup-covid-coverage.aspx

## Acknowledgements

Not applicable

## Supplementary Material

Supplementary material Table 1 is attached.

## Abbreviations

CART: Classification and Regression Tree
CI: Confidence Interval
CIT: Conditional Inference Tree
NaN: Not a Number
OR: Odds Ratio
US: United States
UK: United Kingdom

