## Supplementary Table 1 for "Using conditional inference to quantify interaction effects of socio-demographic covariates of US COVID-19 vaccine hesitancy"

Table 1: Description of participants and breakdown of the COVID-19 vaccine acceptance questions

|  | overall |
| --- | --- |
| n | 16,322 |
| Gender (%) |  |
| Female | 7,429 (45.52) |
| Male | 8,893 (54.48) |
| Household income level (%) |  |
| [0, 36,000) | 2,065 (12.65) |
| [36,000, 60,000) | 2,711 (16.61) |
| [60,000, 90,000) | 3,279 (20.09) |
| [90,000, 120,000) | 3,021 (18.51) |
| [120,000, 180,000) | 2,847 (17.44) |
| [180,000, 240,000) | 1,143 (7.00) |
| [240,000,∞) | 1,256 (7.70) |
| Education level (%) |  |
| Less than high school | 59 (0.36) |
| High school | 1,313 (8.04) |
| Bachelor’s degree | 3,885 (23.80) |
| Postgraduate degree | 4,129 (25.30) |
| Other | 6,936 (42.49) |
| Age group in years (%) |  |
| 18-24 | 82 (0.50) |
| 25-54 | 5,685 (34.83) |
| 55-64 | 3,844 (23.55) |
| 65+ | 6,711 (41.12) |
| Party (%) |  |
| Democrat | 6,822 (41.80) |
| Republican | 4,105 (25.15) |
| Independent | 5,002 (30.65) |
| Other | 393 (2.41) |
| Ethnicity (%) |  |
| White | 14,318 (87.72) |
| Asian | 270 (1.65) |
| Black | 717 (4.39) |
| Hispanic | 836 (5.12) |
| Other | 181 (1.11) |
| Employment status (%) |  |
| Full-time | 6,932 (42.47) |
| Part-time | 1,393 (8.53) |
| Involuntary-unemployed | 606 (3.71) |
| Not-in-labor-force | 7,391 (45.28) |
| Trust in government (%) |  |
| Yes | 5,703 (34.94) |
| No | 10,619 (65.06) |
| Accept COVID-19 vaccine if generally available (%) |  |
| Yes | 11,616 (71.17) |
| No | 4,706 (28.83) |
