## Supplementary material for "Using conditional inference to quantify interaction effects of socio-demographic covariates of US COVID-19 vaccine hesitancy": IRB abstract and title

*** Full Title of Research Protocol**

**Short Title**

SoDem

**Abstract: Provide a simple explanation of the study and briefly address (in 1 to 2 sentences) each of the following points: rationale; intervention; objectives or purpose; study population or sample characteristics; study methodology; description of study arms (if appropriate); study endpoints or outcomes; follow-up; statistics and plans for analysis.**

--Description and rationale: Vaccine hesitancy is a public health problem that needs to be better understood at the population level. Current studies have not revealed how different socio-demographic covariates interact with vaccine hesitancy (dependent variable).

--Intervention: None

--Objectives or purpose: We use survey data made available by Gallup to understand how different socio-demographic covariates interact with vaccine hesitancy (dependent variable).

--Study population or sample characteristics: representative US population polled by Gallup during the previous 12 months and purchased by us under an academic license

--Study methodology: use regression-based (odds-ratio) and conditional inference mathematical techniques to study correlational relationship between individual-level socio-demographic variables and vaccine hesitancy. Individual PII is not available.

--Data description of study arms (if appropriate): not applicable

--Study endpoints or outcomes: Odd-ratios for various socio-demographic groups, and a conditional inference tree, all showing the relationships between socio-demographic variables and vaccine hesitancy.

--Follow-up: none

--Statistics and plans for analysis: Odds ratios will be reported with confidence intervals, and conditional inference tree will only be built for variables that are significant at p-value levels of 1% or less.
