## Supplementary material for "Using conditional inference to quantify interaction effects of socio-demographic covariates of US COVID-19 vaccine hesitancy": IRB submitted protocol

**Study Title:** Measuring socio-demographic covariates of COVID-19 vaccine hesitancy

**PI Name:** Mayank Kejriwal

**Study Procedures**

1. Background/Rationale (previously iStar section 11 (exempt) or 12 (expedited/full board))

COVID-19 vaccine hesitancy has become a major issue in the U.S. as vaccine supply has outstripped demand and vaccination rates slow down. At least one recent global survey has sought to study the covariates of vaccine acceptance, but an inferential model that makes simultaneous use of several socio-demographic variables has been lacking. Several studies have highlighted key factors associated with vaccine hesitancy in different countries, and considerable heterogeneity has been observed. We are interested in the US-context for our study. An influential international survey that provides a template for our own analysis is provided in the footnote on the next page where we describe our methods. Our study is expected to contribute to the discourse on vaccine hesitancy in light of the COVID-19 pandemic, using the most recent Gallup data, but from a distinctly social science and demographic lens.

2. Purpose/Objectives/Aims/Research Questions (previously iStar section 11 (exempt) or 12 (expedited/full board))

The objective of this study is to understand the strength and direction of correlational relationships in the US between a standard set of sociodemographic variables such as age group, household income, ethnicity, employment status and several others, and vaccine hesitancy as measured through the response to a Gallup survey question. We hope that these findings will help us to better understand vaccine hesitancy at a national level.

3. Participants (sample) (Consistent with iStar sections 10 (expedited and full board) and 22 (for all submission types)
   i. The Gallup Panel is a probability-based, nationally representative panel of U.S. adults. Members are randomly selected using random-digit-dial phone interviews that cover landline and cellphones and address-based sampling methods. The Gallup Panel is not an opt-in panel. Gallup weights the obtained samples each day to adjust for the probability of select and to correct for nonresponse bias. Nonresponse adjustments are made by adjusting the sample to match the national demographics of gender, age, race, Hispanic ethnicity, education and region. Demographic weighting targets are based on the most recent Current Population Survey figures or the aged-18-and-older U.S. population.
   ii. Participants do not include children and do not specifically target any members of a special population
4. Recruitment/Screening Process (sampling strategy)

This project only involves secondary analysis of data that has already been collected by Gallup, and we do not recruit, sample or screen participants ourselves. Per Gallup (see attached document), “the COVID-19 web survey began fielding on March 13, 2020 with daily random samples of U.S. adults, aged 18 and older who are members of the Gallup Panel. Approximately 1,200 daily completes were collected from March 13 through April 26, 2020. From April 27 to August 16, 2020 approximately 500 daily completes are being collected. Starting August 17, 2020, the survey moved from daily surveying to a survey conducted one time per month over a two week field period (typically the last two weeks of the month).”

5. Methods (previously iStar sections 9, 12, or 13, 16, 19, 20, and 21 depending on the type of study/application)
   a. Detailed description/methodology of the project:
      i. Step-by-step methods:
         A. As a first step, we will process socio-demographic variables and vaccine hesitancy data as a table and use R for our analysis. Gallup already provides data using a fairly standard form; hence, we do not expect to do data standardization beyond importing the files into a program.
         B. Next, we will use univariate regressions and odds-ratios to understand the strength and directions of relationships between sociodemographic variables and vaccine hesitancy, in a very similar vein to the odds-ratio analysis of Lazarus et al. 2020^[1]^.
         C. We will report all odds ratio with confidence intervals, including ones that are non-significant so that there is no reported bias in our results.
         D. We will also explore and analyze vaccine hesitancy using a classification conditional inference tree (CIT) model (which uses unbiased recursive partitioning) at the individual level. CITs embed tree-structured regression models into a well-defined theory of conditional inference procedures. It is unnecessary to consider data imputation techniques, since the models generate high prediction performance (namely, accuracy of 75.5%). The R package *party* will be used for our recursive partitioning analysis, with the max depth set to 4, and with the p-value required to be less than 0.01, in order for a node to be split by the model.
         E. Odds-ratio and conditional inference tree methods are inspired by similar analysis in a COVID-19 international survey study by Lazarus et al. as well as another study^[2]^ in Nature Climate Change published a few years ago.
      ii. Our study does not include any intervention by the researchers since we are only analyzing secondary data that has already been collected by Gallup
      iii. For expedited and full board studies only: Not applicable as we are asking for exempt IRB.
      iv. Data/specimens are anonymous i.e. Not labeled with any personal identifying information (e.g., name, email address, IP address) or a code that the research team can link to personal identifying information.
      v. All data will be stored and processed locally on work-provided laptops and desktops only within the PI’s group. Data will be deleted upon conclusion of the study and publication of the results.
      vi. We plan to publish all research findings in peer-reviewed scientific venues, possibly preceded by publishing in a pre-print server.
   b. Instrumentation
      i. Questionnaires/Survey Measures (names and citations)
         A. Survey measures were not developed by the PI but by Gallup, which is an expert in conducting this kind of survey. I have attached a document that summarizes their methodology and codebook.
      ii. Qualitative instruments
          A. We will not use any open-ended questions (even if they were administered by Gallup and are included in the survey) in our study.
   c. Data Analysis
      i. Data analysis will be conducted by studying the odds ratios and their implications, along with 95% confidence intervals. We will also study the conditional inference trees and their implications. In structure, the analysis will be very similar to the one conducted in the international survey by Lazarus et al. 2020 (see footnote) published last year.
      ii. *If conducting secondary data analysis from a data set obtained from an existing data source, upload the data collection form(s) identifying the variables to be captured and analyzed in iStar section 13:* we have uploaded the Gallup COVID-19 survey methodology and codebook, which contains complete details on the variables.

1. Lazarus, J. V. et al. A global survey of potential acceptance of a covid-19 vaccine. Nat. medicine 27, 225–228 (2021).
2. Lee, T. M., Markowitz, E. M., Howe, P. D., Ko, C.-Y. & Leiserowitz, A. A. Predictors of public climate change awareness and risk perception around the world. *Nat. climate change* **5**, 1014–1020 (2015).
