## Supplementary material for "Using conditional inference to quantify interaction effects of socio-demographic covariates of US COVID-19 vaccine hesitancy": IRB exempt approval notice

### FW: Study Approval Notice Sent

---

**From:** <>
**Sent:** Wednesday, September 29, 2021 12:27:46 PM (UTC-08:00) Pacific Time (US & Canada)
**To:** Mayank Kejriwal <>
**Subject:** Study Approval Notice Sent

USC

University of Southern California Institutional Review Board
1640 Marengo Street, Suite 700
Los Angeles, California 90033-9269


| | |
|---|---|
| Date: | Sep 29, 2021, 12:27pm |
| Action Taken: | **Approve** |
| Principal Investigator: | Mayank Kejriwal<br>INFORMATION SCIENCES INSTITUTE (ISI) |
| Faculty Advisor: | |
| Co-Investigator(s): | |
| Project Title: | **SoDem** |
| Study ID: | **UP-21-00692** |
| Funding: | |

The University of Southern California Institutional Review Board (IRB) designee reviewed your iStar application and attachments on 9/29/2021.

Based on the information submitted for review, this study is determined to be exempt from 45 CFR 46 according to §46.104(d) as category (4).

As research which is considered exempt according to §46.104(d), this project is not subject to requirements for continuing review. You are authorized to conduct this research as approved.

**If there are significant changes that increase the risk to subjects or if the funding has changed, you must submit an amendment to the IRB for review and approval. For other revisions to the application, use the "Send Message to IRB" link.**

**The materials submitted and considered for review of this project included:**
1. iStar application UP-21-00692, dated 8/10/2021
2. Study protocol
3. Gallup COVID-19 survey methodology and codebook

**NOTE TO PI:** For any future revisions to this study or future secondary data studies, you will be required to utilize the Secondary Data Analysis Protocol Template available at https://oprs.usc.edu/forms-and-templates/.

**DATA COLLECTION FORM**

You must use the data collection form submitted.

**STUDY PERSONNEL**

Individuals who are knowledgeable about the protocol must obtain consent from subjects for participation in a study. Specifically, they must be able to describe the purpose, procedures, benefits, risks, and alternatives to participation in the study. They must be able to answer subjects' questions about the protocol and about risks of the research procedures and alternatives. The PI must identify all individuals who will obtain consent and attest that they fit the above criteria. The PI is ultimately responsible for ensuring that ethically and legally valid consent is obtained from all research subjects.

**Funding source(s): N/A – no funding source listed**

Attachments:

Social-behavioral health-related interventions or health-outcome studies must register with **clinicaltrials.gov** or other International Community of Medical Journal Editors (ICMJE) approved registries in order to be published in an ICJME journal. The ICMJE will not accept studies for publication unless the studies are registered prior to enrollment, despite the fact that these studies are not applicable "clinical trials" as defined by the Food and Drug Administration (FDA). For support with registration, go to www.clinicaltrials.gov or contact Jean Chan (, 323-442-2825).

**Important**

The principal investigator for this study is responsible for obtaining all necessary approvals before commencing research. Please be sure that you have satisfied applicable requirements, for example conflicts of interest, bio safety, radiation safety, biorepositories, credentialing, data security, sponsor approval, **clinicaltrials.gov** or school approval. IRB approval does not convey approval to commence research in the event that other requirements have not been satisfied.

This is an auto-generated email. Please do not respond directly to this message using the "reply" address. A response sent in this manner cannot be answered. If you have further questions, please contact iStar Support at (323) 276-2238 or.
